# Machine Learning to Predict Neonatal Mortality Using Public Health Data from São Paulo - Brazil

**DOI:** 10.1101/2020.06.19.20112953

**Authors:** Carlos Eduardo Beluzo, Luciana Correia Alves, Everton Silva, Rodrigo Bresan, Natália Arruda, Tiago Carvalho

## Abstract

Infant mortality is one of the most important socioeconomic and health quality indicators in the world. In Brazil, neonatal mortality accounts to 70% of the infant mortality. Despite its importance, neonatal mortality shows increasing signals, which causes concerns about the necessity of efficient and effective methods able to help reducing it. In this paper a new approach is proposed to classify newborns that may be susceptible to neonatal mortality by applying supervised machine learning methods on public health features. The approach is evaluated in a sample of 15,858 records extracted from SPNeoDeath dataset, which were created on this paper, from SINASC and SIM databases from São Paulo city (Brazil) for this paper intent. As a results an average AUC of 0.96 was achieved in classifying samples as susceptible to death or not with SVM, XGBoost, Logistic Regression and Random Forests machine learning algorithms. Furthermore the SHAP method was used to understand the features that mostly influenced the algorithms output.

## 1 Introduction

Infant Mortality (IM) is an important measure of health in a population as a crude indicator of the poverty and socioeconomic level. It also shows the availability and quality of health services and medical technology in a specific region. Comparisons of the Infant Mortality Rate (IMR), which is presented as the deaths of children less than one year old per 1,000 alive births, are used in needs assessments and to evaluate the influence of public policies. IM is categorized as Neonatal, when the death occurs after postpartum and until 28 days of life; and as the Post-Neonatal, when the death occurs from 29 days of life until one year of life.

Neonatal Mortality Rate (NMR) and IMR are therefore very important indexes of public health and development level of a country. Actions to decrease NMR and IMR result in the improvement of infant mortality and survival, which can positively influence the national public situation of health [8]. The neonatal mortality accounts to approximately 60% of the IM in developing countries [44]. This dimension of IM is important because, from the point of view of the World Health Organization (WHO) [37] and United Nations Children’s Fund (UNICEF) [45], the first month of life is the period in which the child is more vulnerable. In Brazil it’s been observed an increase in the participation of neonatal mortality in the total of IM cases over more than one decade, as we can see in Figure 1.

**Fig 1:**
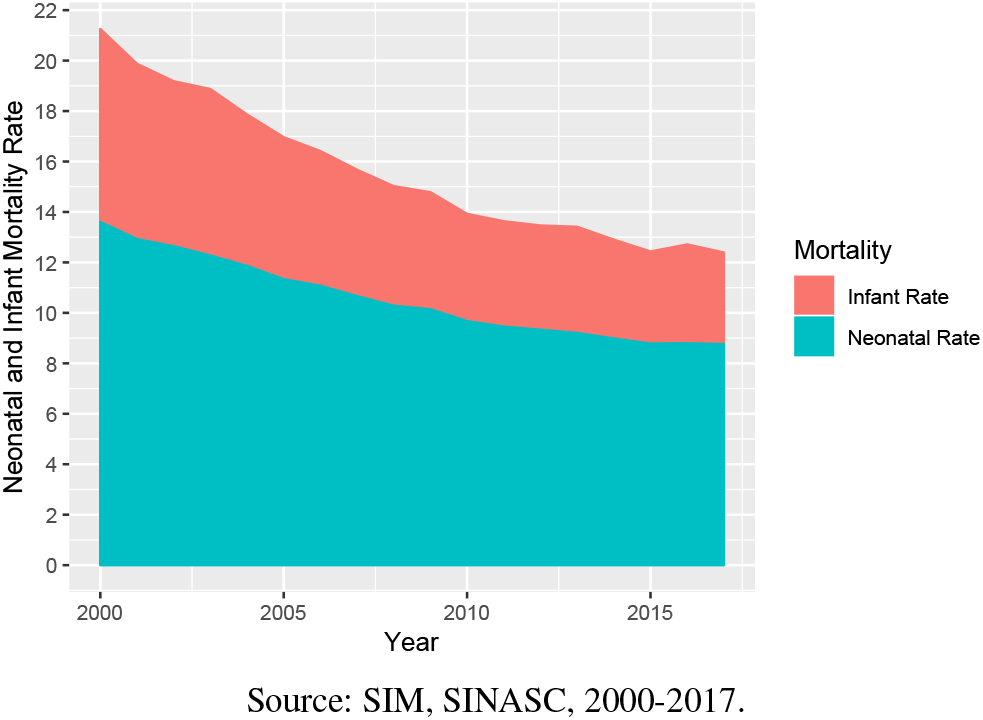
Participation of Neonatal Mortality in the Infant Mortality - Brazil.

Child Mortality is an worldwide concern in public health as defined by the United Nation (UN) on the global development goals when setting the reduction of the infant mortality until 2015 as a target. Brazil achieved this Millennium Development Goal, but national rates do not reveal the persistent inequalities remaining between geographic regions and population groups. Regions and populations with lower incomes are at greater risk of infant deaths. In addition to the disparities arising from socioeconomic and geographic factors, infants in the first week of life (early neonatal death) did not reduce satisfactorily and now represent the greatest challenge to the advancement of addressing infant mortality in the country [42].

The problem of infant mortality in Brazil has become relevant, since the available data and their respective analyzes point out to the persistence of disparities between regions, states and populations with different socioeconomic characteristics, despite the constant tendency of general decline [42]. Besides that, evaluating the data from the period of 2015 to 2017, it can be observed a reverse behavior of the neonatal mortality in Brazil, which after more than 20 years of decline, the NMR started to raise, as illustrated on Figure 2. Moreover, in Brazil, in 2017, we had a NMR of 9 deaths per 1,000 live births, while in developed countries, the NMR is on average 4 neonatal deaths per 1,000 live births.

**Fig 2:**
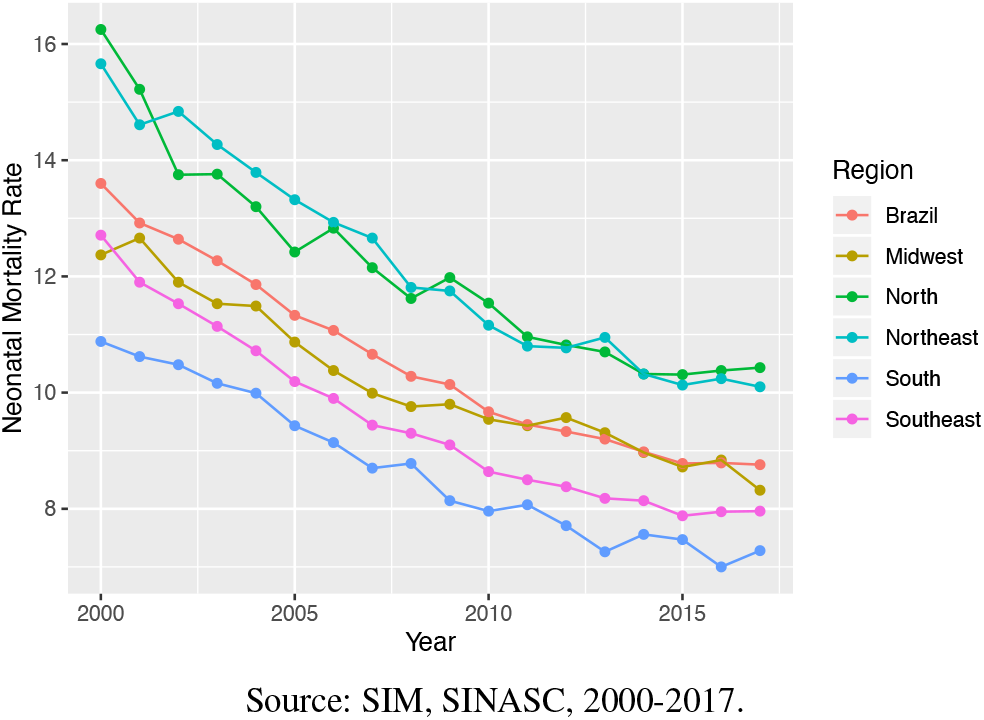
Neonatal Mortality by Macroregion and Total of Brazil.

In Brazil, since the enactment of the Federal Constitution of 1988, a large part of the burden of coping with neonatal mortality has been imposed on municipalities, that have adopted a prominent position in the implementation of public health policies [22, 3]. Sã o Paulo, the capital city of the state of Sã o Paulo, located in the southeast of Brazil, has the lowest NMR, which was 7.5 per 1,000 live births in 2017.

The factors associated with neonatal mortality are deeply articulated and influenced by the maternal and newborn biological characteristics, social conditions and the care provided by the health services [32, 11]. In 2003, Mosley and Chen [31] proposed a hierarchical model based on the hypothesis that socioeconomic factors determine behaviors, which, in turn, have an impact on a set of biological factors. According to their model, biological factors are those directly responsible for death. The hierarchical model brings a great advance to the development of public policies, since information coming from studies that are limited to only a group of risk factors result in inadequate recommendations to assess the deaths among children, as they present a limited vision of the phenomenon.

Similarly, on this paper it is proposed the use of this kind of features but along with machine learning supervised methods to asses neonatal death risk, as well as to identify the features that mostly influence it. **On this sense, the base of our scientific hypotheses is: different characteristics of the mother and the newborn as maternal obstetrics, related to the newborn and to the care assistance on prenatal and delivery are able to predict neonatal mortality more than only socioeconomic characteristics of the mother**.

### 1.1 Machine Learning Applied to Demographic Research

Most of the demographic studies in Brazil search for specific factors related to infant and neonatal mortality, based on the use of descriptive analysis like spatial analyzes, multiple statistical and logistic parametric regression and, in general, using small datasets.

Nascimento et. al. [32] proposed an hierarchical model to analyse a dataset with 264 neonatal deaths while Migoto et. al. [29] uses a dataset containing 157,604 live births and 903 early neonatal deaths (up to the sixth day of life). Both works found some strongly related factors to total neonatal mortality and early neonatal mortality. It was observed that neonatal deaths were related mainly to the quality of the prenatal care and direct care labor. These features were measured through some variables such as: number of prenatal consultations, type of labor, professional responsible for the childbirth (doctor on call, obstetrician, nurse or other).In addition, some associations were found regarding the reproductive history of the mother, such as if the mother presented fetal losses in previous pregnancies. They also identify some relation with presence of malformation and to maternal socioeconomic conditions (mother’s education, marital status and mother’s race).

Migoto et. al. [29] also points that maternal age indicates a higher chance of early neonatal death among adolescent mothers and those who were 35 or older when compared to mothers who were 20 to 34 years old. Regarding the education, the mothers of children who died before completing one week of life studied until they were seven. Moreover, the children of women who had no partner were more likely to die when compared to women who were married.

While aforementioned works relay on traditional statistics methods, machine learning (ML) approaches start to be highlighted among some international works. Nguyen [33] proposes to use machine learning approaches to analyze in-hospital child mortality using as features the final diagnosis, for example, children with meningitis or malnutrition diagnosis were most likely to die. This work had the concern to make models that were capable of detecting the death in the sample since the outcome was a rare event.

Pan [38] observed that ML models were capable of identifying more then 150 high risk pregnant women besides the paper-based risk assessment already used in the social services in Illinois. So, the ML methods were capable of improving the efficiency of decision making and improvements in the identification of high-risk pregnancy eligible to receiving specific health services.

The ML approach proposed by Podda et. al. [40] aimed at estimating the survival of newborns prematurely and compared machine learning models with the most common logistic methods in these types of analyzes. Author’s methods predict the survival of preterm neonates better than logistic models and, thus, allow a better approach for identifying risks and allowing the improvement of decision quality and identification of risks. They used Neural Networks and observed that although logistic regression models and other linear models are more easily understood and interpretative and their results are easily used as risk measures, this ease of interpretation is lost when interactions between variables are present, and in this case, neural networks can take into account interaction between variables and non-linearities with the variable outcome.

Hsieh et. al. [16] proposed a comparison of ML models with the aim to predict the mortality of patients with unplanned extubation in intensive care units. They observed that even with limited data points (341), they were able to develop a good prediction model. This authors worked with an unbalanced data, with the Random Forest model presenting best recall and precision values compared to the others models used in that work (Support Vector Machine, Artificial Neural Networks, Logistic Regression Model).

According to the best of our knowledge, there are no studies in Brazil analyzing the risk of neonatal mortality with machine learning methods. Furthermore, there are no reported results for this kind of problem using as input features from Mortality Information System (Sistema de Informaçã o de Mortalidade - SIM) and the National Information System on Live Births (Sistema de Informaçã o de Nascidos Vivos - SINASC).

**The main contributions of this work are: (1) proposition of a new method, modeled as a classification problem, based on ML approaches, to detect neonatal death; (2) a comparative study to assess efficacy of different types of machine learning classifiers; (3) an analysis of important features to improve problem understanding**.

The rest of this work is divided as following: Section 2 presents details of the methodology including classification algorithms description and experiments protocol defined for this paper, as well as the dataset construction process and respective exploratory analysis. Section 3 brings details of performed experiments and respective results, including results of the feature importance method. Finally in Section 4 it is presented a discussion of the experiments results, the main conclusions and perspectives for future works.

## 2 Methodology

The methodology proposed on this paper ^1^, depicts on Figure 3, follows three main steps:

**Fig 3:**
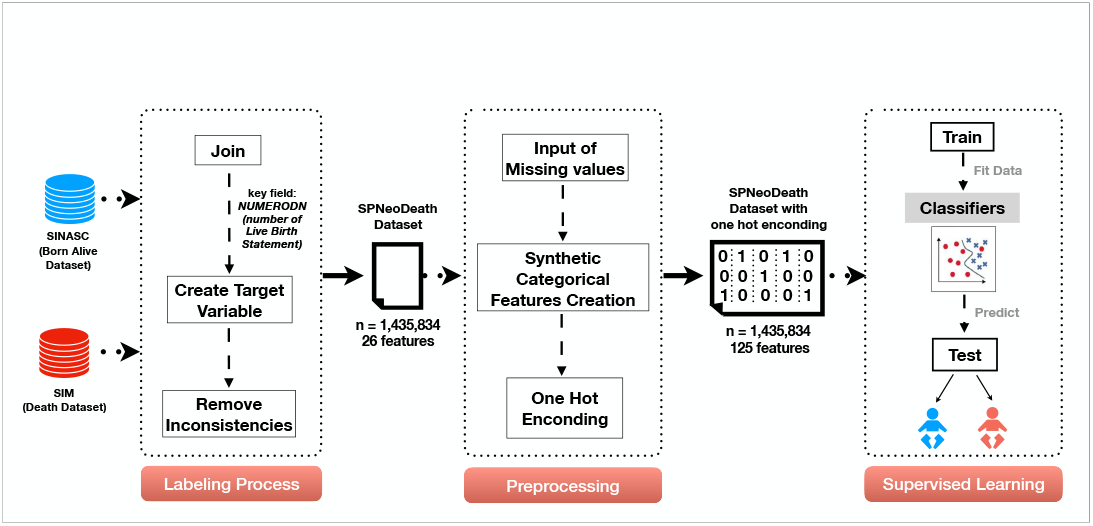
Overview of the methodology followed on this paper.

- **Labeling Process** consists in performing a joined analyzes between samples from SINASC and SIM datasets; creating the target variables; and removing records having any kind of data inconsistency;
- **Data Preprocessing:** in this third step common techniques were applied in order to prepare the dataset to be used with selected ML methods: (i) fill missing values; (ii) transform all features in categorical; and (iii) transform the data set by using the *One Hot Encoding* technique;
- **Supervised Learning:** finally, in the last step, four ML methods were experimented *Support Vector Machines, Logistic Regression, Extreme Gradient Boosting, and Random Forests*. Additionally to evaluating the features that mostly influence algorithm results, the *Shapley Additive Explanation* method was applied. A brief introduction of all these methods is presented in the next section.

### 2.1 Labeling Process

Data used in this study was extracted from birth and death records in the City of Sã o Paulo between 2012 and 2018. Also, we just keep death records from neonatal period (when the child died before the first 28 days of life). The municipality of Sã o Paulo is the one with better data quality and the data was collected directly from the Municipal Health Office of São Paulo (SMS Secretaria Municipal de Saúde de São Paulo).

Although São Paulo has the best quality and best levels of neonatal rate when compared to the rest of Brazil, as depicted in Figure 4, these events occurred in heterogeneous ways with smaller or even lower reductions in the most vulnerable populations, as the reflection of unfavorable life conditions of the population, health care and socioeconomic inequalities [12].

**Fig 4:**
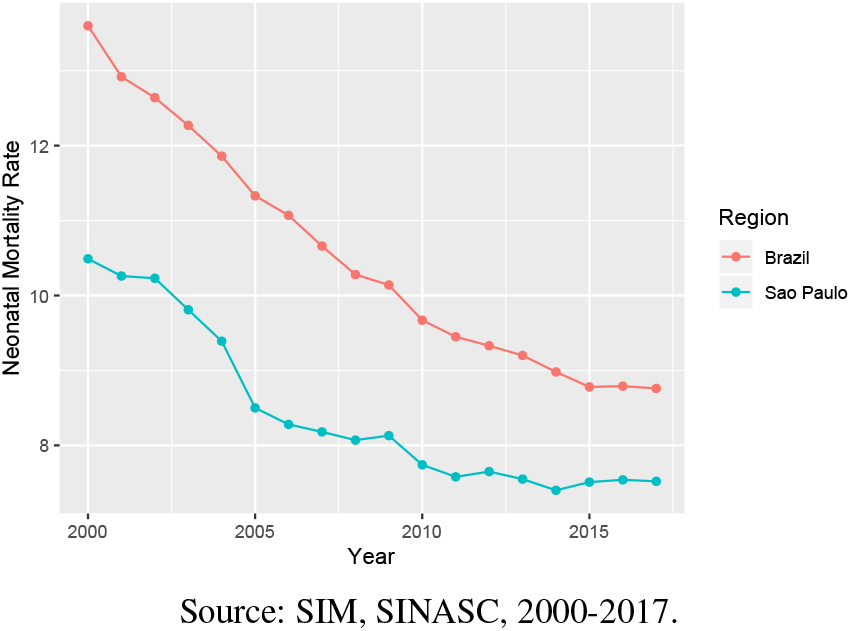
Comparison of Neonatal Mortality by São Paulo and Total of Brazil: 2000 to 2017.

#### 2.1.1 SINASC and SIM Data Sources

The data used on this paper was extracted from SINASC and SIM, that are the two main sources of information on births and deaths in Brazil respectively. SINASC is fed based on the Live Birth Statement (Declaraçã o de Nascido Vivo - DNV) [36], and was used to retrieve demographic and epidemiological data on the infant, mother, prenatal care and childbirth. SIM has the main goal of supporting the collection, storage and management process of death records, in Brazil [30], and was used to label death records on SINASC, using DNV as an association key.

DNV is a standard document prepared by the Ministry of Health (Ministério da Saúde - MS) and mandatory throughout the national territory for the registration of a child’s birth. It must be filled in all live births, whatever the circumstances of the birth: hospitals, maternity, emergency, household, public roads, vehicles, etc. Similarly, we have the Death Certificate (Declaraçã o de Ó bito - DO) that is the document used to collect information about mortality and it is used as the basis for the calculation of vital statistics, such as the calculation of the Brazilian neonatal mortality rate.

Even though filling out the DNV and the DO is mandatory, there is a significant deficiency in data quality due to many situations as losses when sending them from hospitals to the city health office’s (which is responsible to report to the MS); fields filled with incorrect values; unknowing information by the person answering, etc.

SINASC and SIM data sets are not initially linked, so to associate birth and death records, a simple union between the datasets was performed by using the DNV field, which is present inn both datasets, as mentioned before. This approach was based on previous papers such as [24, 43].

After the union, a new field was added in the resultant data set, to label the samples as being a neonatal death (deaths occurred before the first 28 days of life) or not. This was achieved by calculating the difference between the birth date (from SINASC) and the death date (from SIM). As in the proposed method the SIM data is used just to allow data set labeling, after this process all SIM fields were removed from the final dataset.

Finally, rows having fields with inconsistent values were removed, and as a result we got a dataset containing 1,435,834 million records, with 24 features (and the target variable) referred on this paper as **SPNeoDeath** dataset. The features are described on Table 1.

**Table 1:**
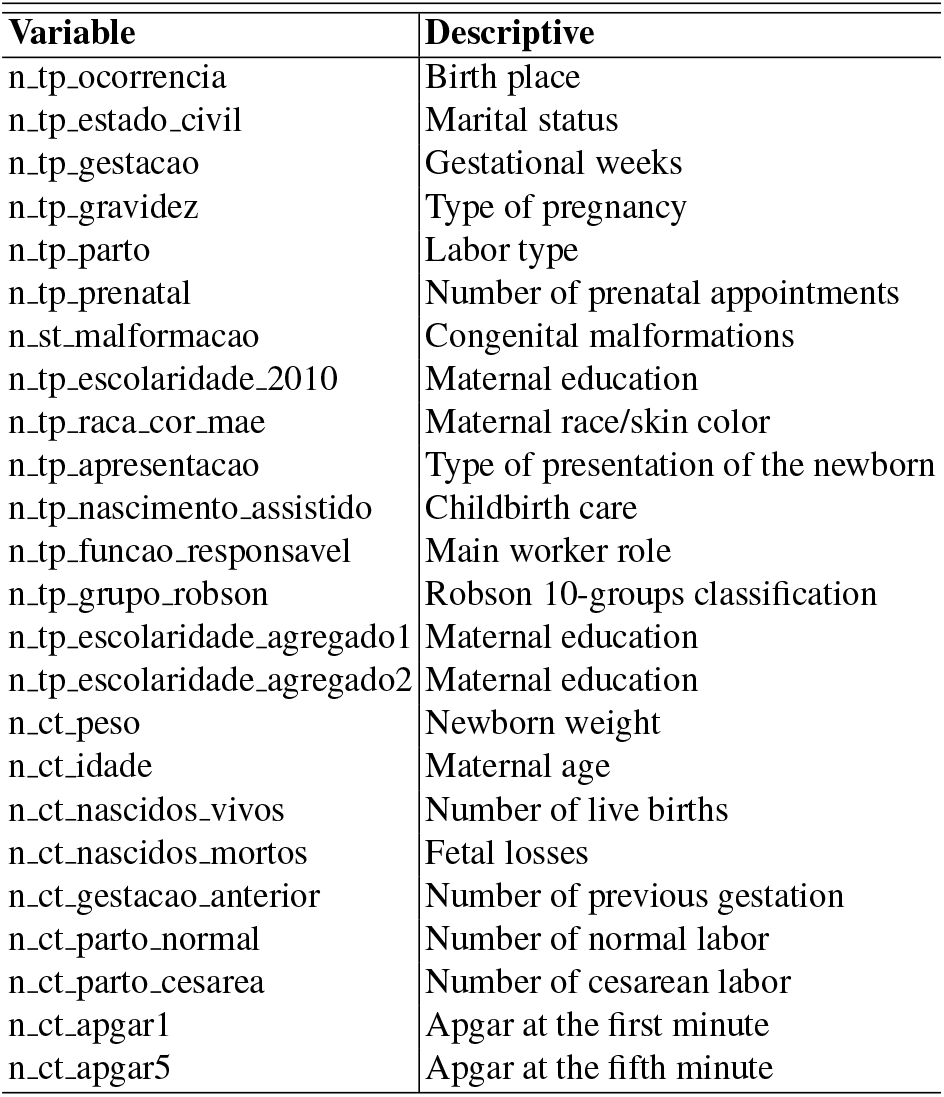
Dictionary of data - variables used in the models

#### 2.1.2 SPNeoDeath: Dataset Description and Exploratory Analysis

From the problem perspective (neonatal mortality) it is important to highlight some specificities regarding possible values for features in the dataset. In order to better understand the problem and the **SPNeoDeath** dataset itself, in this section we provide some descriptions of the dataset as well as the results of an exploratory analysis.

The exploratory analysis presents the values of the features acordding to its distribution (%) of demographic, socioeconomic, maternal obstetrics, related to newborn and previous care characteristics in São Paulo from 2012 to 2018. Information is presented in the categories: (a) socioeconomic maternal conditions features; (b) maternal obstetrics features; (c) newborn related features; and (d) previous care related features. Furthermore, some statistics regarding the features values distribution among target classes are also presented (deaths/lives).

1. **Socioeconomic maternal conditions features:** age, education, marital status and skin color. Most of the mothers were between 15 and 34 years old, but the age was predominantly 25 to 29 years old (24%); 54% were either married or in a stable relationship; 56% of the mothers had had 4 to 7 years of education; and 57% were white. The complete values distribution is on Figure 5.

**Fig 5:**
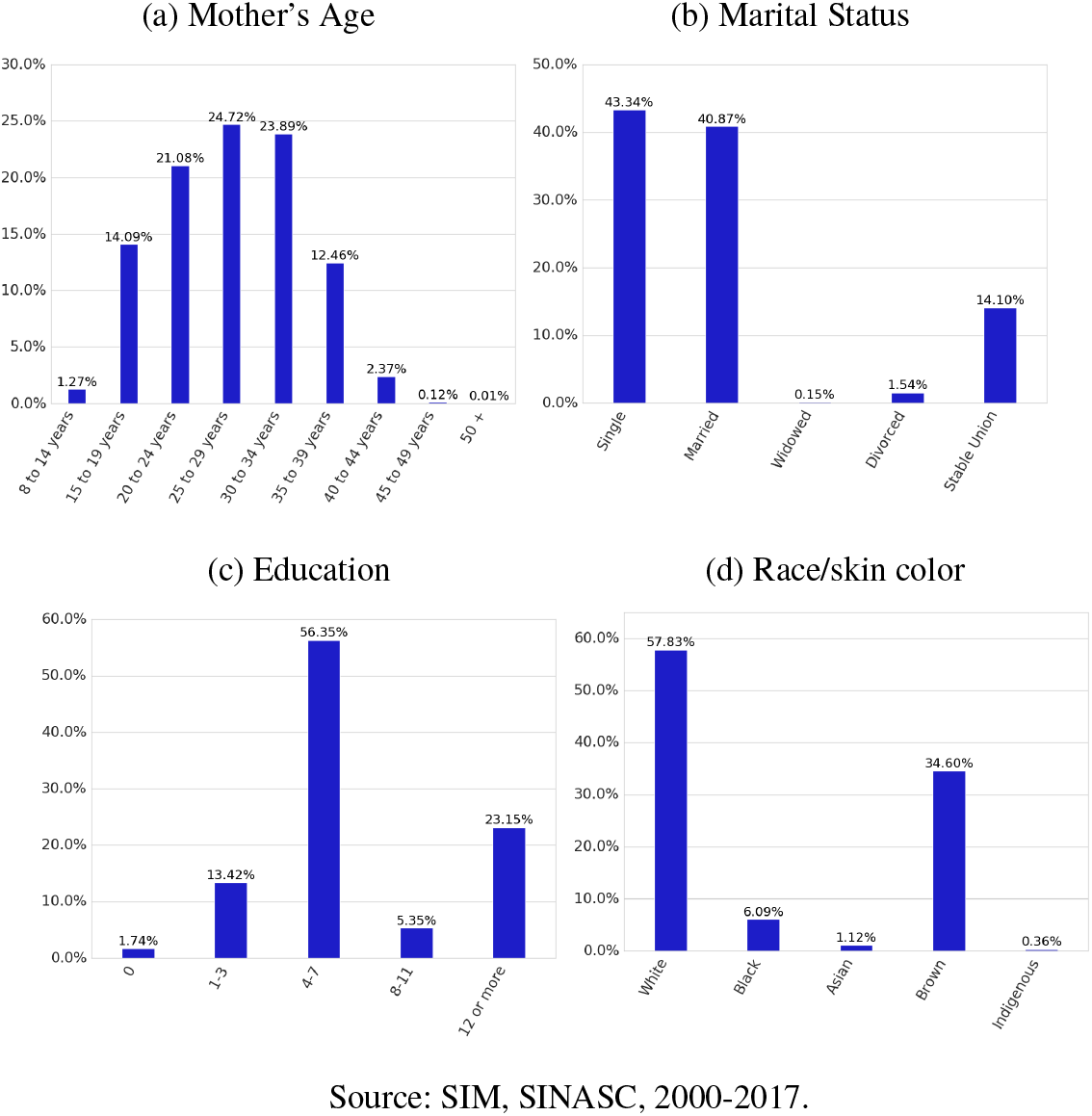
Socioeconomic Maternal conditions
2. **Maternal obstetrics features:** number of live births, fetal losses, number of previous pregnancies, number of normal and cesarean labor, estimate type and type of pregnancy. Most of the mothers had 0 to 3 children (98%); about 99%, 97%, 99%, and 99% had 0 to 3 fetal losses, previous pregnancies, normal labor and cesarean labor, respectively; approximately 97% had a single pregnancy and 87% had 37 to 41 gestational weeks. The complete values distribution is on Figure 6.

**Fig 6:**
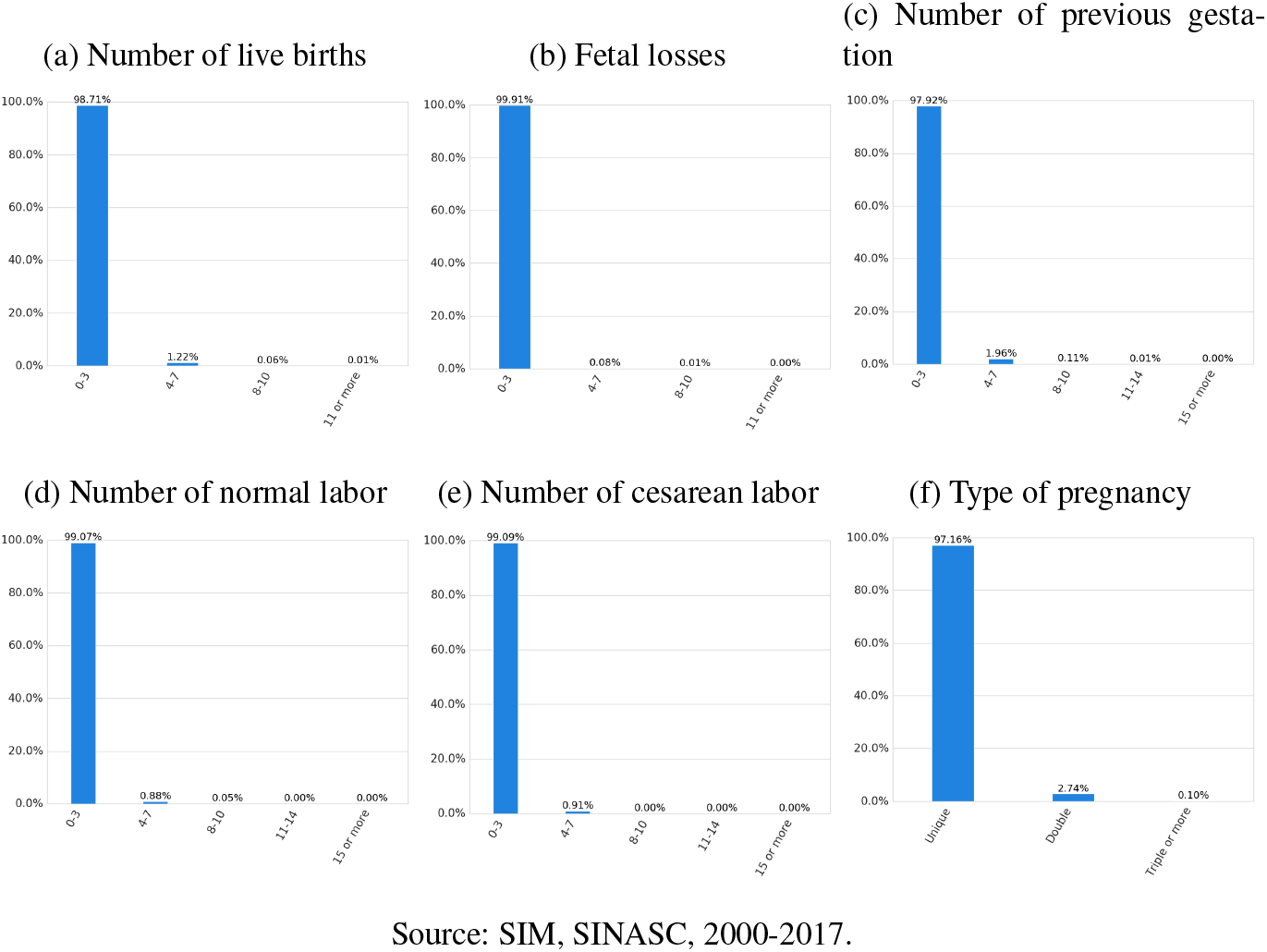
Maternal Obstetrics/Previous care
3. **Features related to the newborn:** weight, number of pregnancy weeks, apgar index first minute, apgar index fifth minute, congenital anomaly and type of presentation of the newborn. Regarding the newborn features, 61% was born weighing between 3,000 and 3,999 grams and 24% between 2,500 and 2,999 grams. Most of the newborns scored 8 to 10 on Apgar in the 1st and 5th minutes (65% and 93%, respectively). The complete values distribution is on Figure 7.

**Fig 7:**
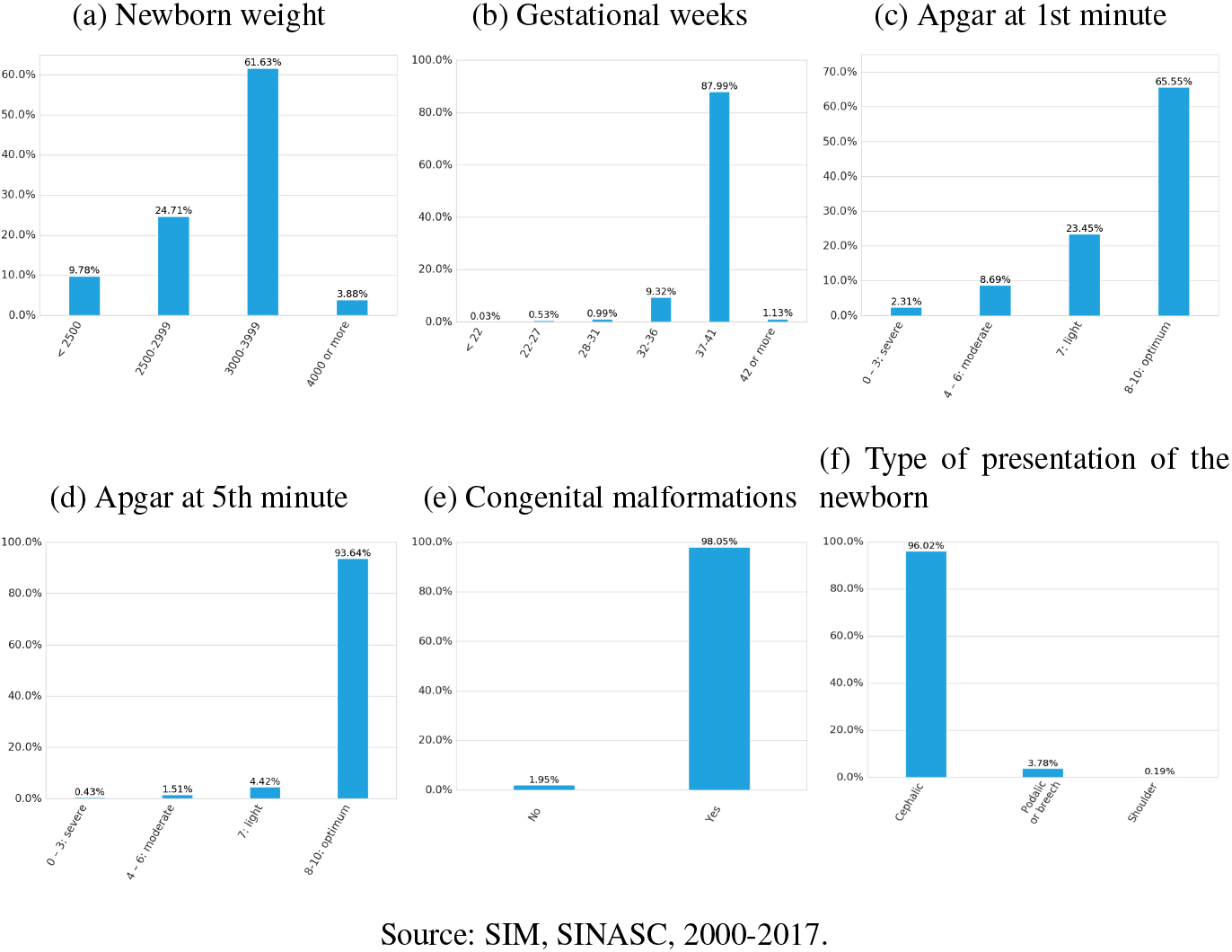
Related to Newborn Characteristics
4. **Features related to previous care:** number of prenatal consultations, labor type, if the cesarean section occurred before labor began, if the labor was induced in the Robson 10-groups classification. Finally, most mother’s have been to more than 7 prenatal appointments (78%); 56% had cesarean labor; 81% had childbirth care for the doctor; and 34% were in Robson Classification Group 2. The complete values distribution is on Figure 8.

**Fig 8:**
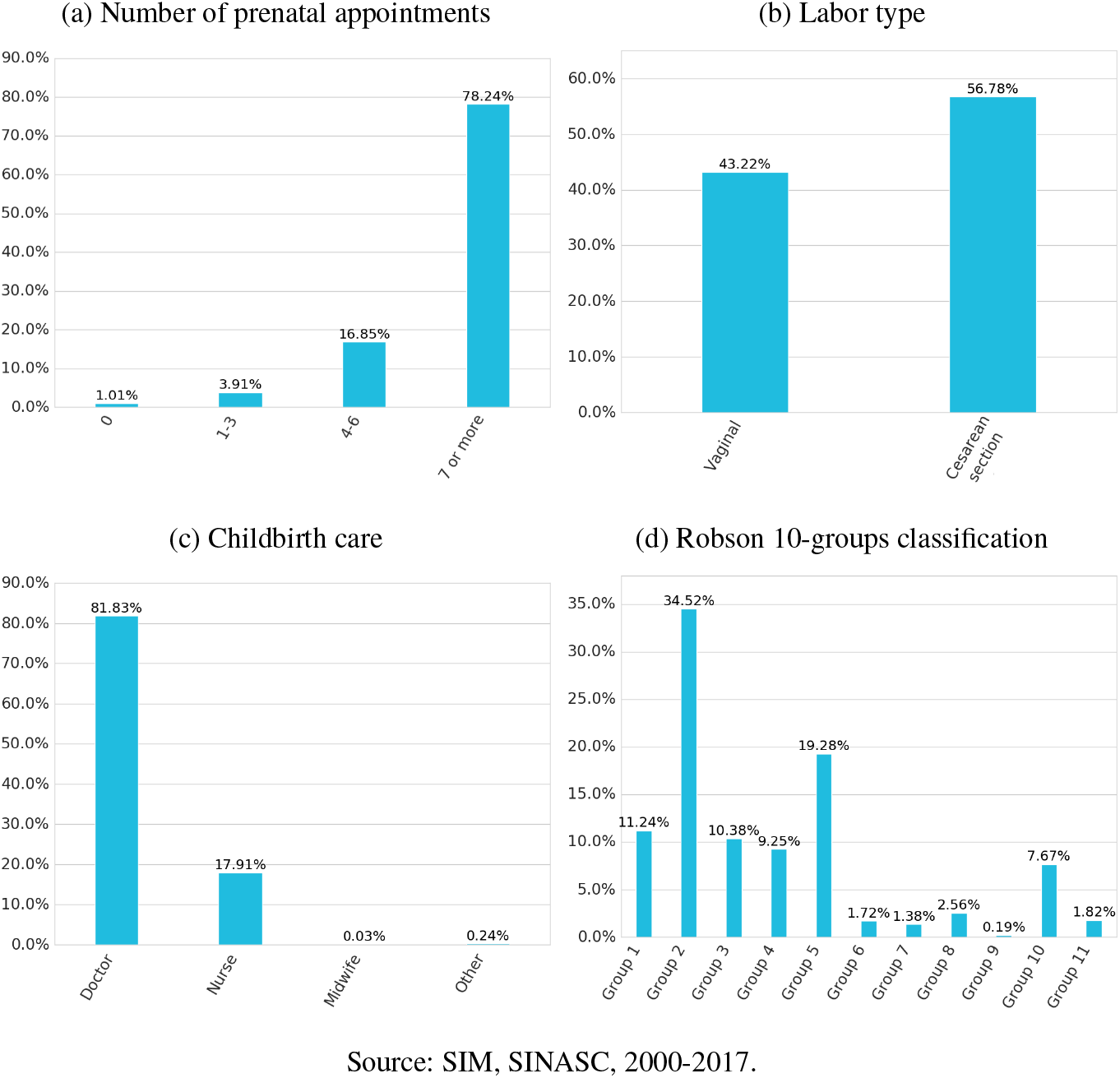
Related to previous care

#### 2.1.3 Features Values Distribution Among Target Classes

##### Sex, age and prenatal consultations

from all neonatal death data 55% was male and 45% was female. The average age of the survivors was about 9 days. Related to the number of prenatal consultations, 38% went to 4 to 6 consultations, 36.5% went more than 7 times and almost 20% went only 1 to 3 times. Observing only those newborn who survived (7,928 samples), we have 78.39% of mothers went to more than 7 consultations and only 1% of those mothers didn’t go to any one, as depicted in Figure 9.

**Fig 9:**
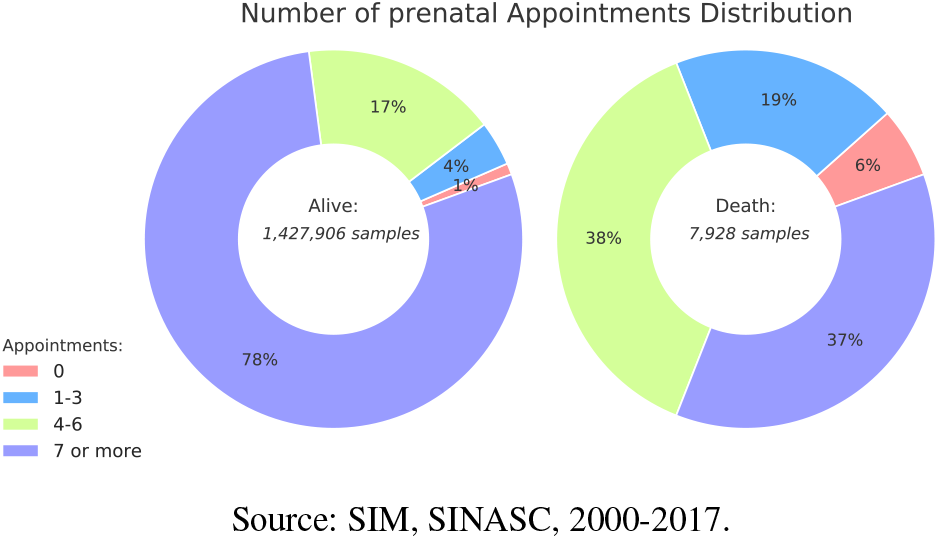
Comparison of Prenatal consultation between death and alive target variable

##### Birth weigth, apgar index and gestational weeks

when analyzing birth weight, which frequency is depicted at Figure 10, 78.75% of newborns who died before the 28th day of life (death class) had insufficient weight - below 2,500 grams, whereas in the alive class only 9.32% were underweight and 91% were born with weight greater than 2,500 grams.

**Fig 10:**
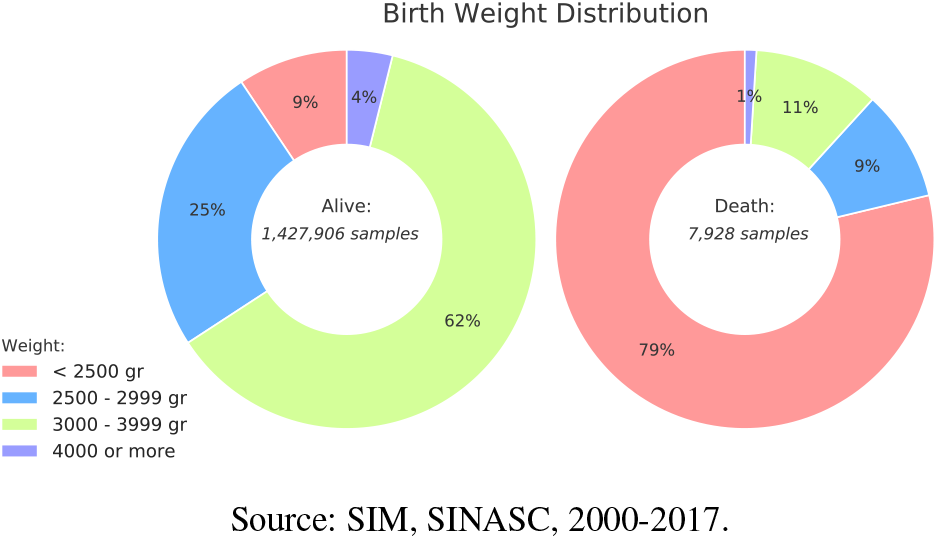
Comparison of birth weight between death and alive classes.

The apgar index (apgar1 and apgar5) are tests done on the newborn just after birth in the 1st minute and then on the 5th minute which assess his general condition and vitality. Related to apgar1, we can observe on Figure 11, in death class, that more than 78% of the newborns in the death class had an index considered serious or moderately severe. In contrast, 63% of the newborns who did not die had an optimal assessment in the first minute of life. Furthermore, Figure 12 depicts results for apgar5, exposing that 51% of the death class had a severe or moderately severe evaluation, while 94.1% of the alive class had an optimal evaluation.

**Fig 11:**
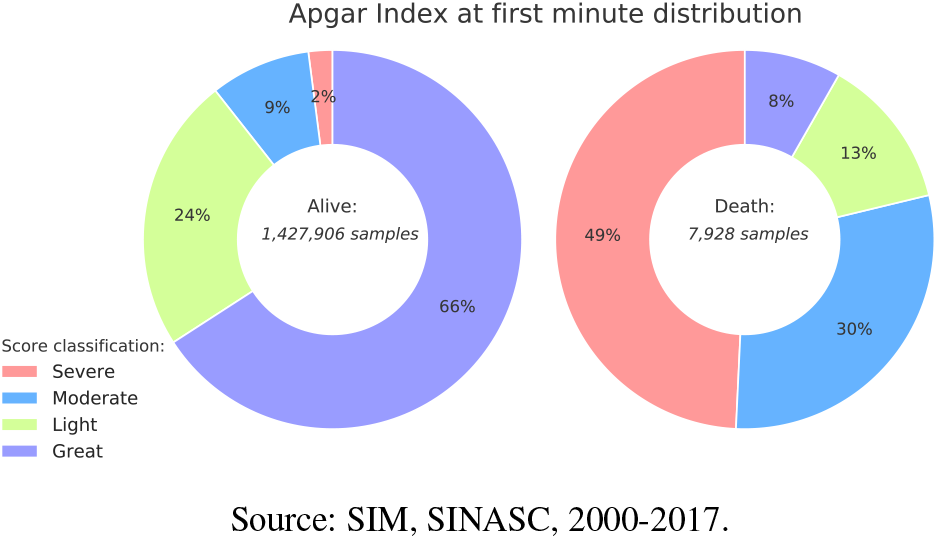
Comparison of Apgar Index at 1st minute between death and alive classes.

**Fig 12:**
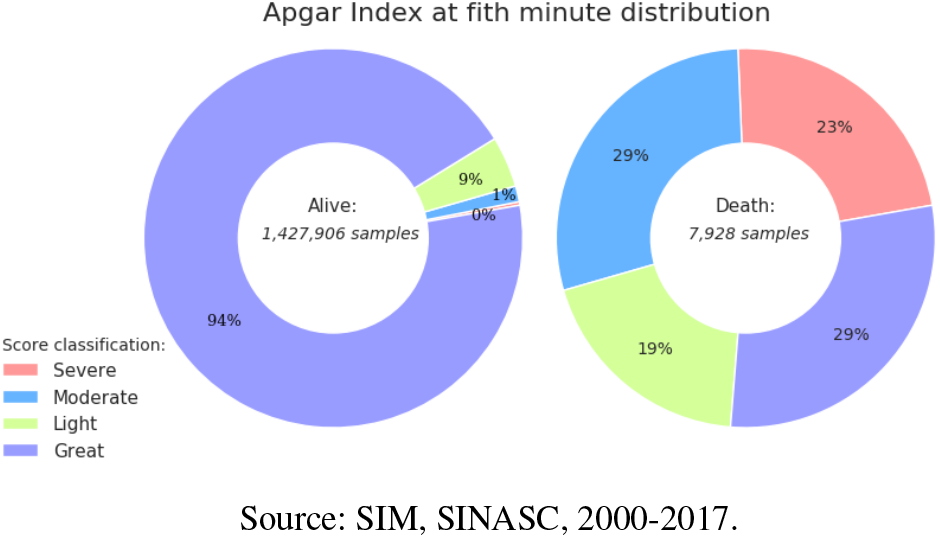
Comparison of Apgar Index at 5th minute between death and alive classes.

Regarding gestational weeks, we can see in Figure 13 considering the death class, that 76.41 % were born preterm, that is, before the thirty-sixth week of gestation. In the alive class, 88.21% were born in the range of 37 to 41 weeks of gestation.

**Fig 13:**
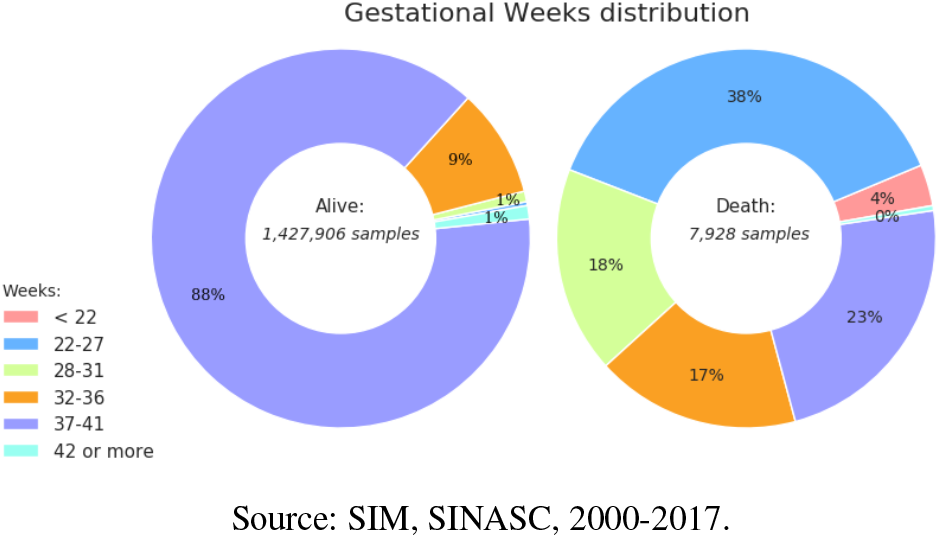
Comparison of Gestational Weeks death and alive classes.

### 2.2 Preprocessing

In a way to achieve best performance in chosen ML classifiers, some common data preprocessing techniques were applied on SPNeoDataset: (i) missing values treatment; (ii) synthetic categorical transformation; and (iii) One Hot Encoding.

#### 2.2.1 Missing Data Processing

In the context of Brazilian public health data, occurrence of missing or inconsistent data is usual and it mostly happens due to the incorrect filling of handwritten forms. In order to take care of this problem, a general approach for demographic studies, based on approaches used in other studies was applied. [34, 25]. Basically, two different techniques were applied: for non-categorical features, with continuous numerical values, the main value for this feature in the dataset is calculated, and the feature is filled with this value; whereas features with categorical values (discrete values) are filled using the most frequent value for this feature in the dataset.

#### 2.2.2 Synthetic Categorical Transformation

Features in the **SPNeoDeath** dataset are mostly categorical, however, some of them are in continuous domain. The proposed method creates the concept of synthetic categorical feature, which encodes the feature value from a continuous numeric domain into a specific discrete value, values which were defined for this paper, and have meaning for the neonatal death risk classification problem. The features were encoded as following:

- **n ct peso:** grouped in four groups as defined by WHO [39]: Low Weight (less than 2500 gr); Insufficient Weight (2500 to 2999 gr); Adequate Weight (3000 to 3999 gr) and Weight Excess (4000 gr or more).
- **n ct idade:** grouped in 9 groups starting with the lowest ages present in the records (8 to 14 years old) and ending at 50 years or older, always creating groups of five-years as determined by different literature methods based on maternal age [19, 23, 35].
- **n ct nascidos vivos:** the number of children born alive related with mother’s reproductive history was divided in 4 groups starting with the group of 0 to 3 live births, 4 to 7, 8 to 10 and 11 or more.
- **n ct nascidos mortos:** the number of children born dead related to the mother’s reproductive history was divided in 4 groups starting with the group of 0 to 3 live births, 4 to 7, 8 to 10 and 11 or more.
- **n ct apgar1:** this feature is a score in the range from number 0 to number 10 (just using integer numbers) [1]. It has been grouped in 4 groups: 0 to 3 (severe); 4 to 6 (moderate); 7 (light); 8-10 (optimum) [26, 14].
- **n ct apgar5:** this feature follows the same shape as the grouping done by n ct apgar1.
- **n ct parto normal:** the number of each woman’s normal deliveries was grouped in 5 groups: 0 to 3 deliveries; 4 to 7; 8 to 10; 11 to 14; 15 or more pregnancies.
- **n ct parto cesarea:** the number of cesarean sections follows the same grouping configuration as the variable n ct parto normal.
- **n ct gestacao anterior:** the number of each woman’s previous pregnancies was grouped in 5 groups: 0 to 3 pregnancies; 4 to 7; 8 to 10; 11 to 14; 15 or more pregnancies.

#### 2.2.3 One Hot Encode

Adoption of discrete values for categorical columns may pose a problem in the context of ML algorithms, since these values may be misinterpreted in an ordinal manner, showing an hierarchy between the values of that given category. In order to solve this problem, we convert our features into dummy variables using a technique commonly referred as One-hot Encoding [4], which generated a vector of *N* positions for each existent category, where *N* is measured by the amount of unique values of a specific category. Then, one of the columns of the vector is filled with the value of 1 (one), indicating the presence of that given category, where the other positions are filled with 0 (zero).

### 2.3 Supervised Learning Methods

At this section, we present ML algorithms which where selected for the problem showed on this paper. The algorithms choice for this paper come from good results achieved in health problems [40, 16, 38, 33, 17].

#### 2.3.1 Support Vector Machines [9]

Support Vector Machine (SVM), is one of the most common methods applied on supervised classification problems mainly because of its excellent accuracy and generalization properties [40, 16]. The basic concept behind SVM is on the discovery of an hyper-plane that can separated the data into the number of the classes, projecting data in a *M* dimensional space by kernel application.

#### 2.3.2 Logistic Regression [15]

Adopting a logistic function to find a model that better fits the data points, the logistic regression model is used to estimate the probability of a binary event happening based on some previously input data. Once our work is based on a binary classification problem, and due to the simplicity of this method, we choose this algorithm, as also employed previously in other related works [16, 38].

#### 2.3.3 XGBoost [7]

Adoption of gradient boosting machines has been proved to push the limits of processing power for boosted trees algorithms. These techniques have been refined to extract most of the system hardware in order to provide a high quality model. A variant of this model was use in the work of Podda et. al. [40], the Gradient Boosting Machine (GBM), and presented a good performance on predicting preterm infant survivor.

#### 2.3.4 Random Forests [6]

Three-based algorithms, provide an easy comprehension of their outcomes, as well as the interaction between features used on classification. Adoption of random forests rely on the fact that this approach makes usage of multiple trees with a random subset of features for training and testing, thus leading to a higher diversity and more robust predictions. This model has a good performance in different literature approaches [38, 33, 40], including prediction in different contexts of infant mortality outcomes using different features related with the child and the mother.

### 2.4 Evaluation Metrics

In order to assess the quality of our method, as well as to provide a baseline for future works, and given we are working in a binary classification problem where weights for missing answers are different for classes ^2^, classic metrics [5] to analyze the ML models results. Also, the following metrics are the common adoption in other papers that are related to public health analysis using ML methods [41].

Besides that, and as an innovation on studies similar to the proposed here, a method to evaluated the features that are mostly influencing the models results has also been applied to the ML model that achieves the better performance on each round of experiments (detailed in next sections). This approach provides a better interpretation of results and on this paper the **SH**apley **A**dditive ex**P**lanation (SHAP) Values [21] method has been used.

#### 2.4.1 Confusion Matrix, ROC Curve and AUC

##### Confusion Matrix

presents a relation between *true positives* (TP) (death samples correctly classified as death), *true negatives* (TN) (alive samples correctly classified as alive), *false positives* (FP) (alive samples misclassified as death), and *false negatives* (FN) (death samples misclassified as alive).

##### ROC Curve

*Receiver Operating Characteristic Curve* - and the **AUC** - *Area Under the ROC Curve* - has been extensively used as measures for machine learning algorithms because it allows the comparison between the classifiers’ performance across the entire range of class distributions and error costs [20].

The ROC Curve is a coordinate system representation that allows to measure the behavior of a binary classifier system as its discrimination threshold is varied. In the y-axis is plotted the *True Positive Rate* (TPR) values, or *sensitivity*; and in the x-axis is plotted the *False Positive Rate* (FPR) values, or *specificity*. Sensitivity and specificity are calculated as presented on Equations 1 and 2 respectively.

The AUC value represents the integral under ROC Curve, and for the convention the bigger its value, the better the model is evluated.

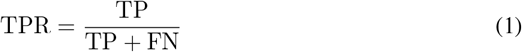

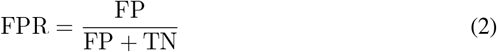

#### 2.4.2 Feature Importance Evaluation with SHAP

Interpretability of machine learning models are a concern across different fields in computer science and applied sciences. For public health and demography this kind of characteristic is specially important. An expert which holds its recommendation using as help a machine learning model, needs to explain and justify the presented conclusions. In this sense, besides the results of the proposed method execution, on this paper it was also applied the SHAP method, to measure features importance and provide a better interpretation of results.

SHAP method belongs to “additive feature attribution methods” which can be simplified by linear function of features. This method tries to come up with a linear regression model for each data point. It replaces each feature (*x*_*i*_) with a binary variable 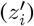 that represents whether *x*_*i*_ its present or not in the model given by:

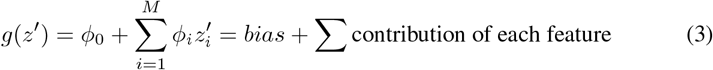

where *g*(*z*^*0*^) is a local surrogate model of the original linear model *f* (*x*) and *c*_*i*_ is how much the presence of the feature *i* contributes to the final output, which helps to interpret the original model.

Formally, a SHAP value measures the influence of a feature *i* to the output *f*_*x*_ produced by a machine learning model by including the feature *i* for all the combinations of features other than *i* defined by:

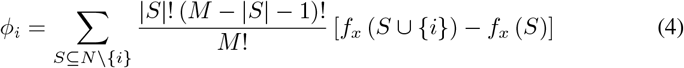

where *S* is the subset of features from all features *N* except for feature 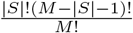 is the weighting factor counting the number of permutations of the subset *S, f*_*x*_ (*S* { *i* }) is the actual output model given all features N (including *i*) and *f*_*x*_ (*S*) is the expected output given the features subset *S*.

In general, the importance is calculated from the reduction of the square error, that is, if this feature was selected in a mode division, in the tree build, and the square error decreased in relation to all the trees. In this paper, the feature importance was calculated by using a Python pre-built library H2O [13].

### 2.5 Experiment Protocols

As already mentioned, since our data contains a substantial imbalance between the classes (neonatal death or alive), models were trained as a priority on balanced data, based on the selection of the same number of records for both classes, resulting in a final dataset with the same amount of records for each class (SPNeoDeath). It was selected a total of 15,858 records, being half of these linked to neonatal death, and the others randomly selected from the subset of non neonatal death records. This approach was chosen due to the fact that most of ML models are better suited for balanced datasets. For benchamark purpose, the same protocols were also performed once using the imbalance dataset, with all 1.4 mi records.

The proposed method was performed in four rounds of experiments. Each round refers to a specific dataset selection and filter approach, but the same four ML models chosen for this paper were tested (SVM, Logistic Regression, XGBoost and Random Forests). At the end of each round, the SHAP method was applied for the ML model showing the best performance (XGBoost achieved the best in all of them, so basically SHAP was applied only to XGBoost).

The experiments were defined according to the subset of features used on that round. As mentioned before, four rounds of experiments were defined, in order to evaluated the influence of having or not a subset of features. The first round was performed using all records and all features, referred here as *imbalance dataset*. The second round was performed using all features, but now with the *balanced dataset* (same number of records for each class, i.e., 7.929 records for the death class, and the same number for alive). The third round was also performed on the balanced dataset, but using only prepartum related features. Finally, in the last round, the experiment was executed on a balanced dataset using only postpartum features.

Regarding the dataset split for training and test tasks when applying ML models (number of folds), for all rounds of experiments, same validation protocols were followed: a 5-folds cross-validation protocol (when the dataset was split into 5 subsets or folds), using 4 folds for training the models, and 1 fold for testing, changing test fold on each interation. This approach is also extensively used on ML models applications.

## 3 Experiments and Results

### 3.1 Environment Setup

The proposed method has been implemented with the Python programming language (3.6), along with the Scikit-Learn (0.21.2), H2O (3.24.0.4), XGBoost (0.90), Pandas (0.24.2) and MatplotLib (3.1) libraries. All the experiments have been performed in a machine with 40 CPU cores, 4 GPU TitanX 12 GB, 120 GB of RAM and 8 TB of storage, running Ubuntu 18.04 (64 bits).

### 3.2 ML Hyperparameters Setup

The experiments were performed using mostly the default parameters of the ML models, and just a few customizations: when using SVM, proposed method takes an *RBF* kernel, along with a *C* value of 1.0 and argument as 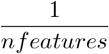, where *n features* is the number of used features; when using Random Forest, the number of estimators were set to 100 and the *criterion* argument as *gini*; XGBoost have been executed using *gbtree* booster with a learning rate of 0.3,a of 0,a *max depth* of 6, and other parameters as default; and Logistic Regression experiments used an *L*2 penalty, a tolerance of 0.0001 and a *C* parameter of 1.0.

### 3.2 Results

#### 3.3.1 Round #1: Imbalanced dataset - All features

For this experiment we evaluated the classification performances about imbalanced dataset composed by all features (prepartum and postpartum features).

The confusion matrix in Figure 14 shows that none of the classification algorithms present a good performance to classify both classes. All classifiers have a good performance to hit samples labeled as alive class. It happens because the dataset is too imbalanced.

**Fig 14:**
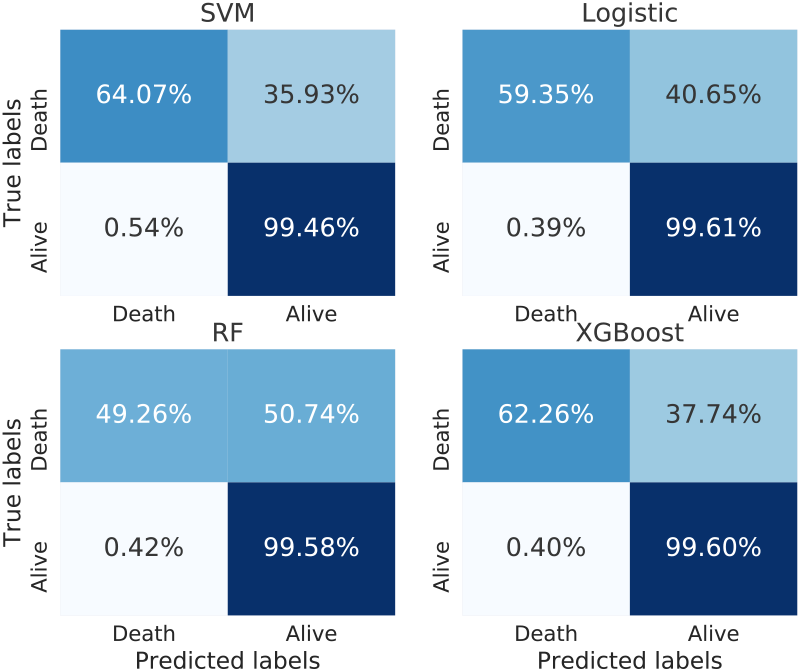
Confusion matrix for imbalanced dataset with prepartum and postpartum features.

Figure 15 depicts associated ROC curves and AUC values for each evaluated classifier. Logistic Regression and XGBoost classification algorithms present a statistical advantage when compared to SVM and RF. Their ROC curves overlap with AUCs values as 0.967 for Logistic Regression and 0.968 for XGBoost.

**Fig 15:**
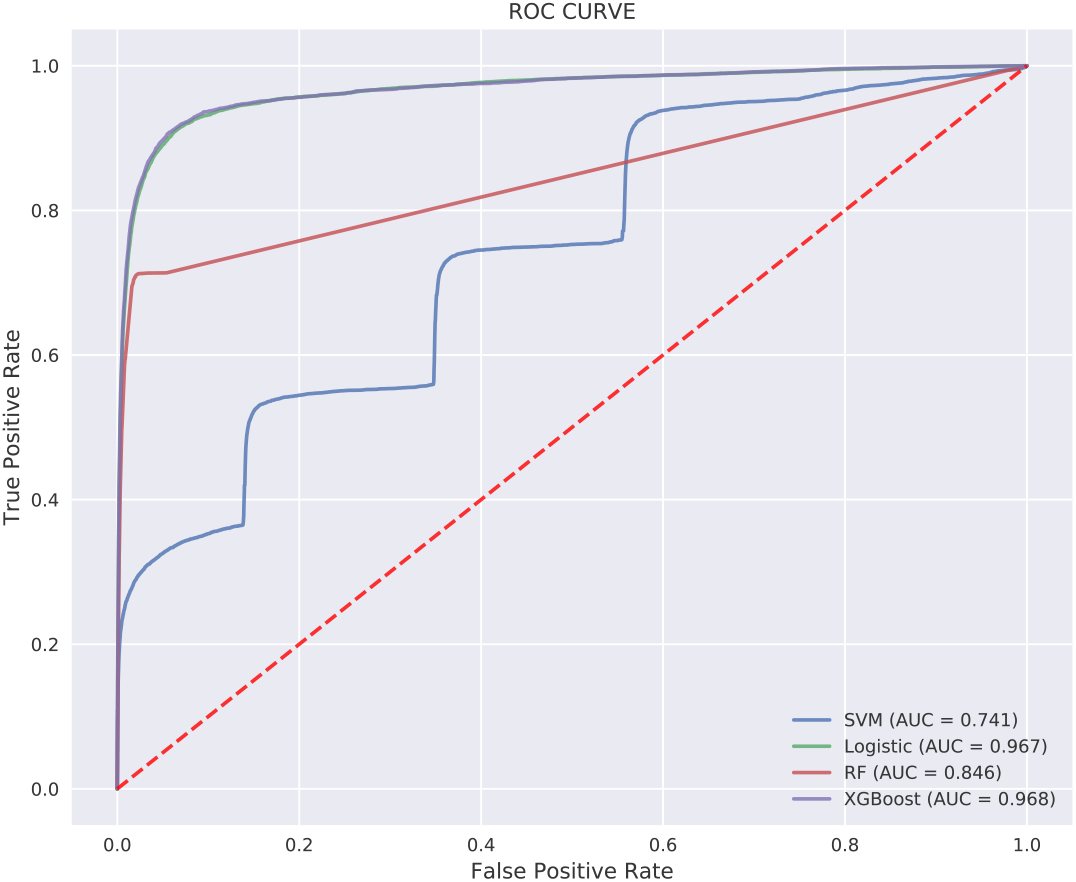
ROC curve for imbalanced dataset with prepartum and postpartum features.

Appling SHAP to explain model output, on this round the top five most relevant features that influence the results are (on this order): apgar at the 1st minute, newborn weight, congenital malformations, apgar at the 5th minute and gestational weeks. The complete feature importance ranking is presented in Figure 16.

**Fig 16:**
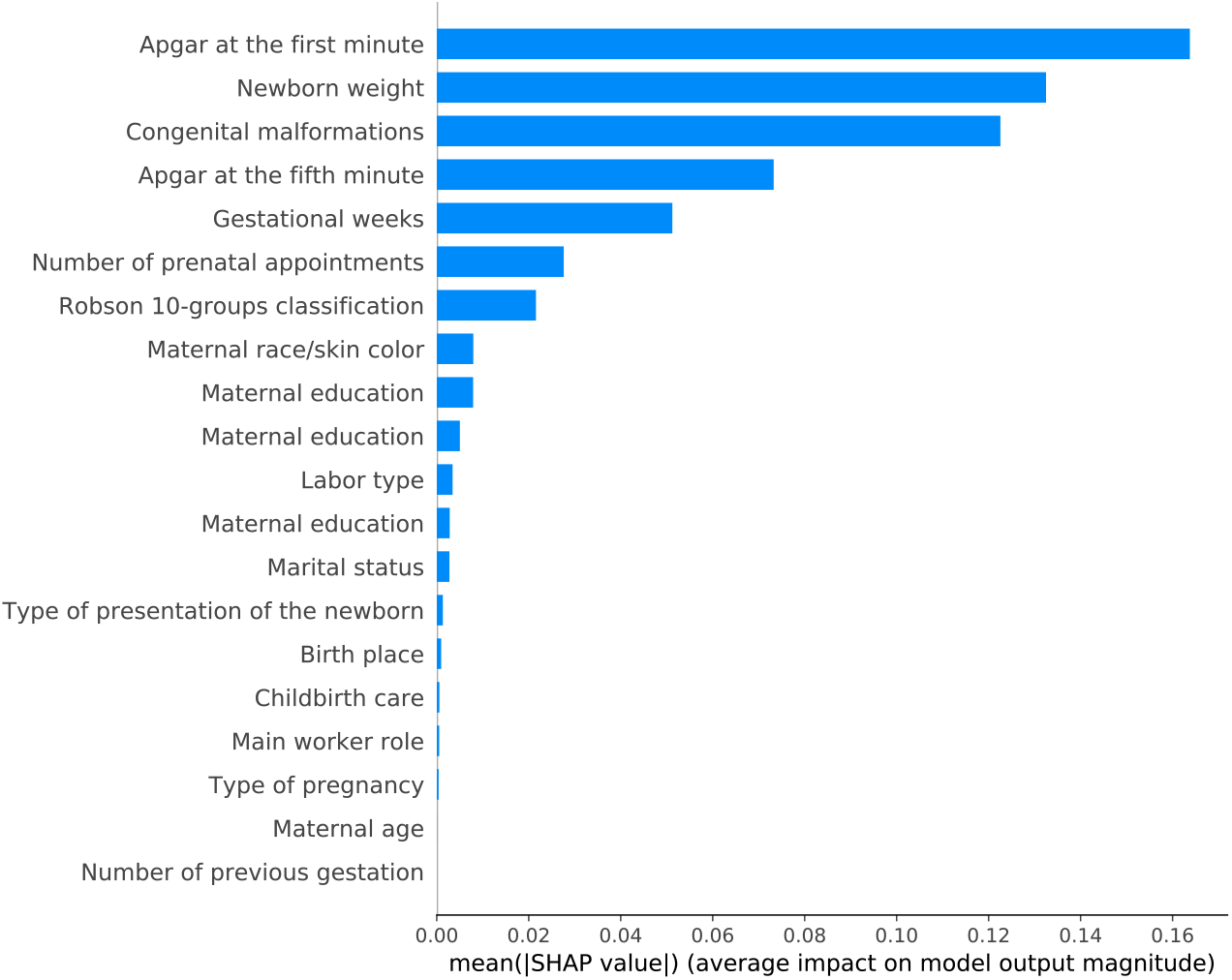
Feature importance for imbalanced dataset with prepartum and postpartum features.

#### 3.3.2 Round #2: Balanced dataset - All features

On this round, the experiment was performed on a balanced dataset composed by all features (next round will be executed with prepartum features only and then with postpartum features only). The performances classification is presented in the Figures 17 and 18.

**Fig 17:**
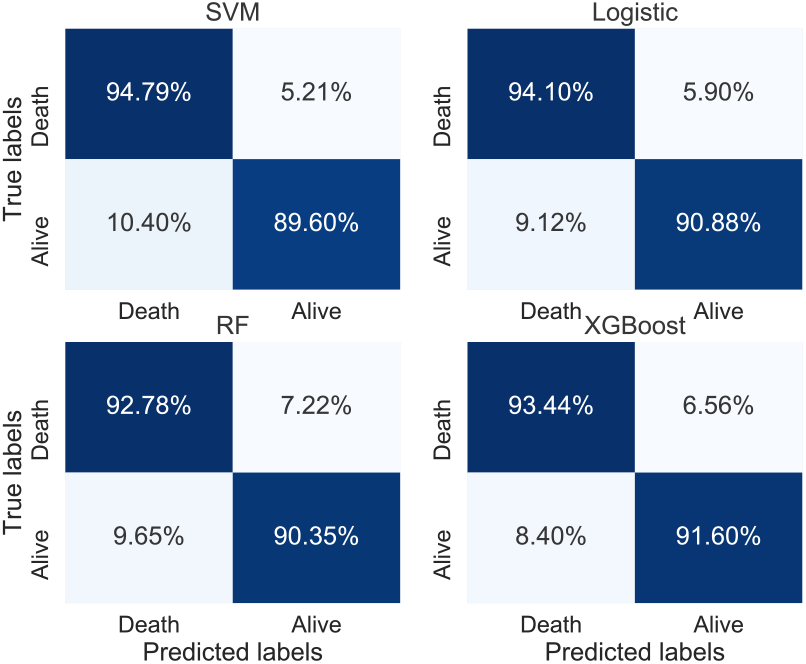
Confusion matrix for balanced dataset with all features.

**Fig 18:**
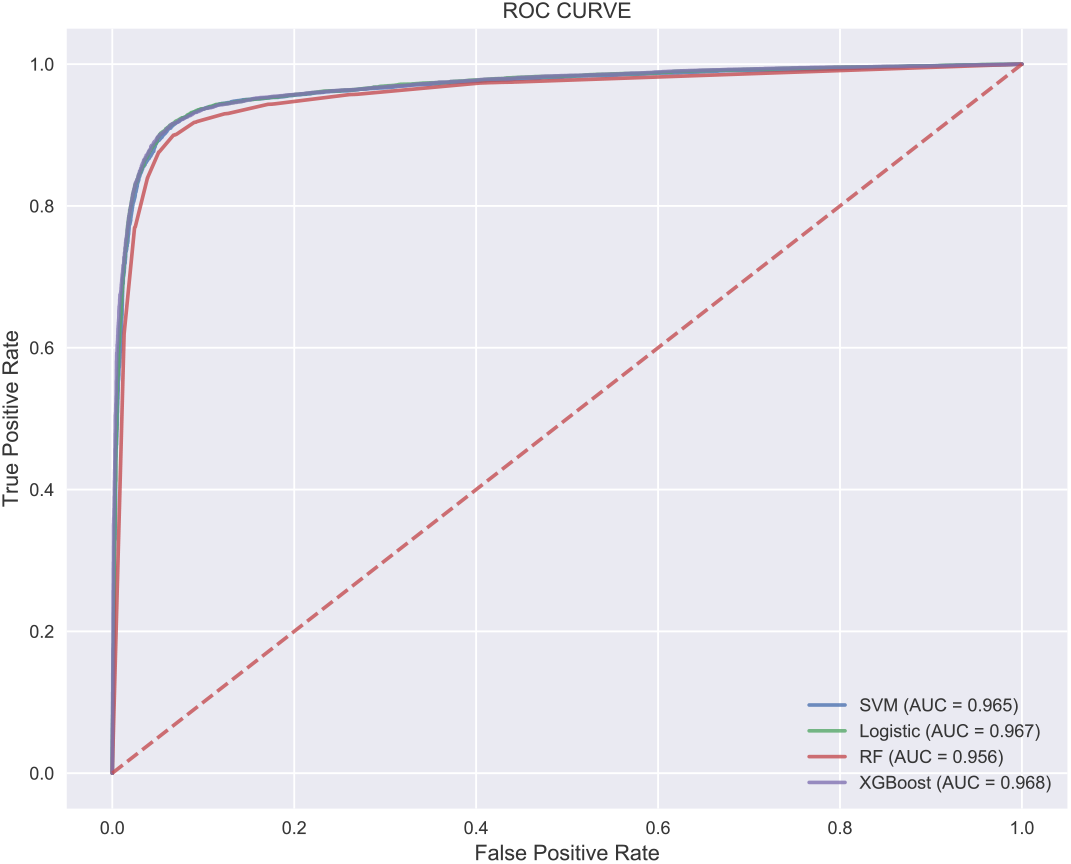
ROC curve for balanced dataset with all features.

From the confusion matrix (Figure 17), we can realize that all classifiers present a good performance to classify both classes. Besides that, the SVM classifier presents a slightly better performance to classify death samples as death (TP), but it does not have a good performance for the alive class.

Analyzing the ROC curve for this scenario, we can observe in Figure 18 that none of the classification algorithms present a statistical advantage when compared among themselves and their AUC values that are around 0.96.

Being able to interpret model output gives a direction of feature engineering, and in this experiment, newborn weight is the feature that most influences the model, followed by congenital malformations, apgar at the 5th minute, apgar at the 1st minute and gestational weeks respectively. Complete feature importance ranking is presented on Figure 19.

**Fig 19:**
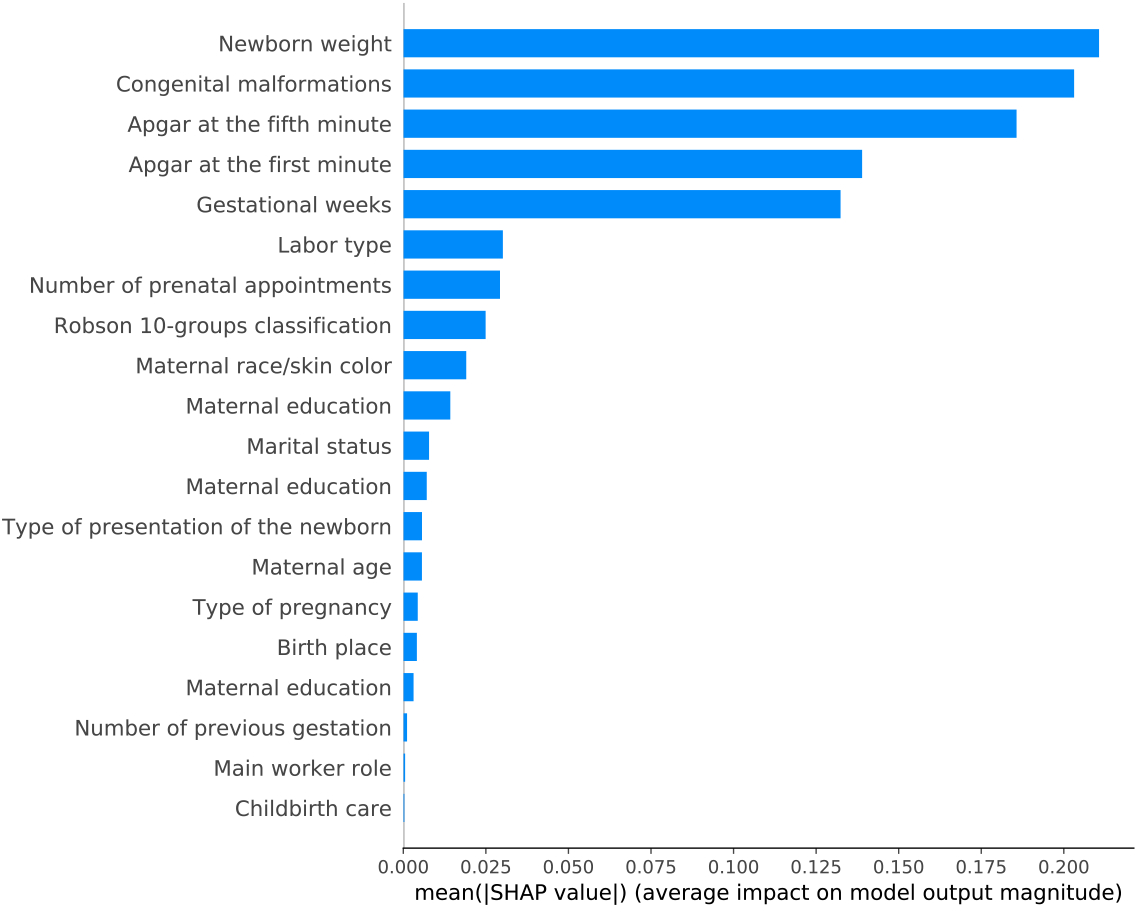
Feature importance for balanced dataset with all features.

#### 3.3.3 Round #3: Balanced dataset - Postpartum features

Round #3 was performed to evaluate the classification performance in a balanced dataset with only postpartum features. Results for this experiment are shown in Figures 20 and 21.

**Fig 20:**
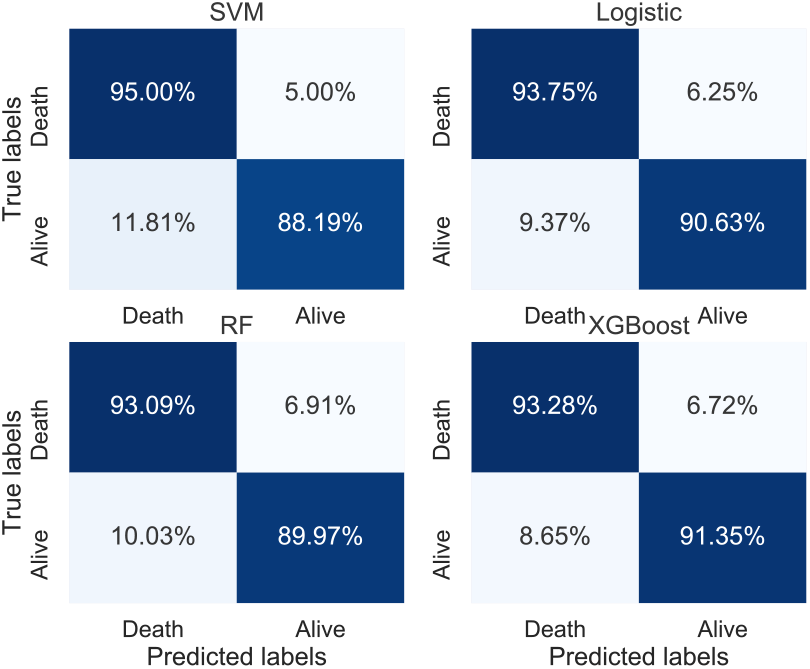
Confusion matrix for balanced dataset with postpartum features only.

**Fig 21:**
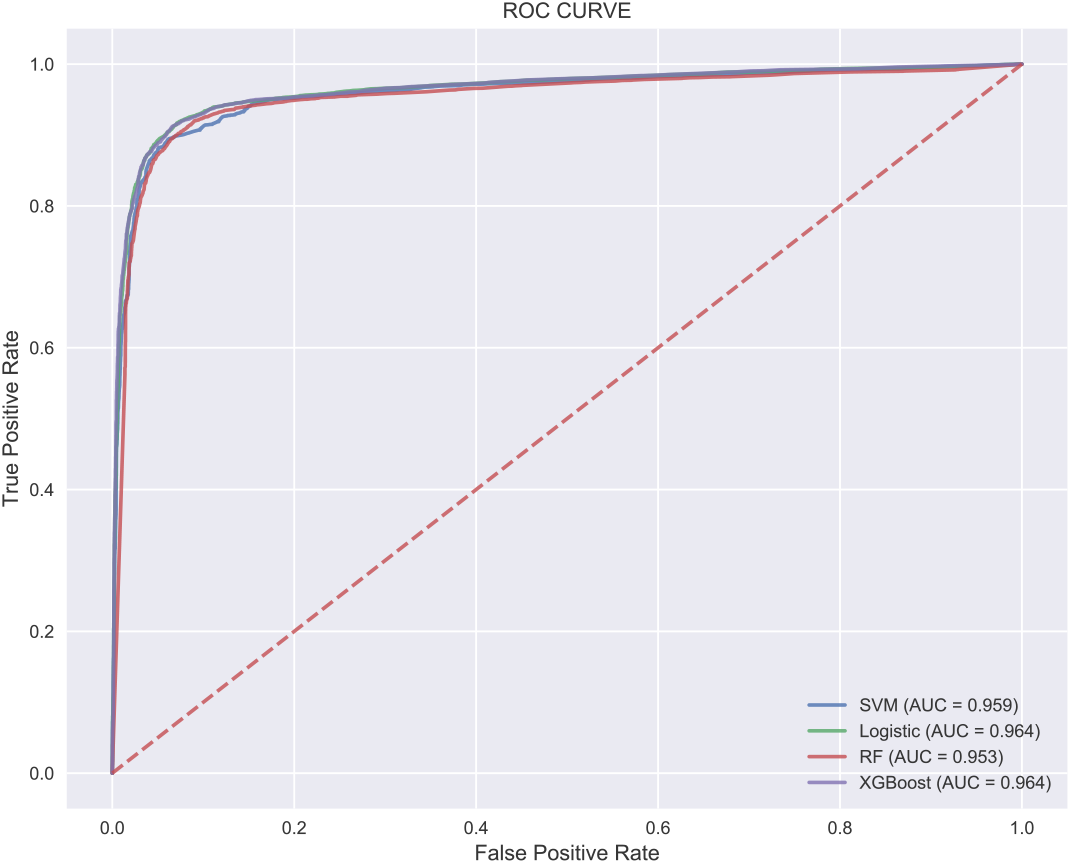
ROC curve for balanced dataset with postpartum features only.

ROC curves and AUC values are very similar to the results present in round #2, where all features were used. So we can conclude that classifiers achieve good predictive performance using only postpartum features. To understand that, Figure 22 shows the importance for each feature performed by using SHAP method. Top five features that impact on model output are: newborn weight, congenital malformations, apgar at the 5th minute, gestational weeks and apgar at the 1st minute.

**Fig 22:**
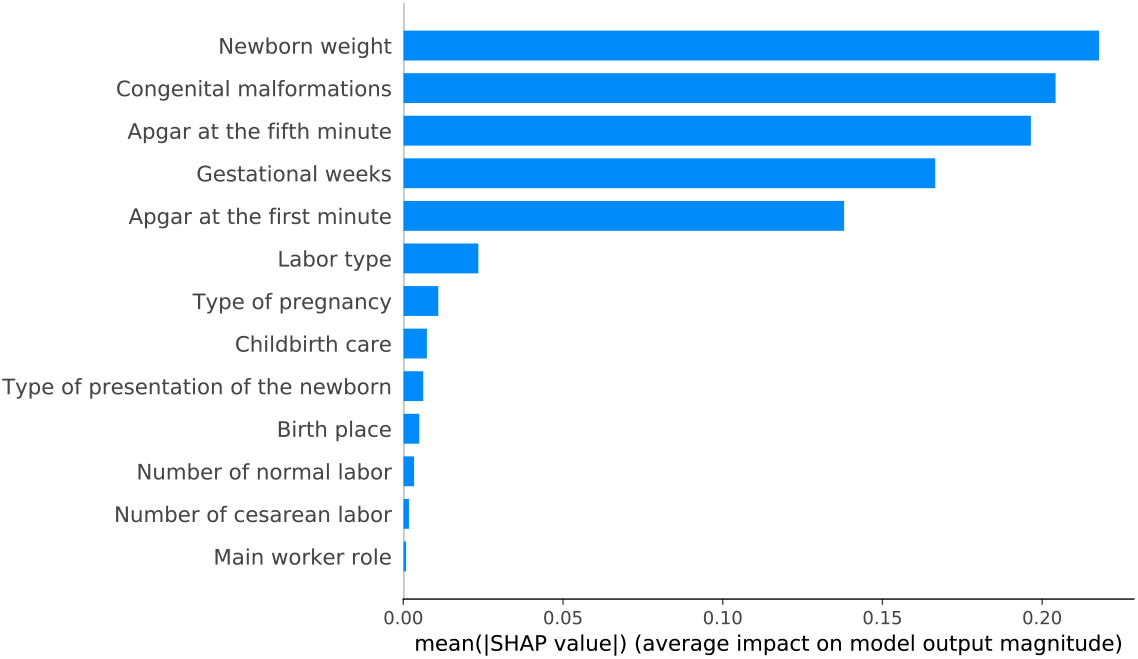
Feature importance for balanced dataset with postpartum features only.

#### 3.3.4 Round #4: Balanced dataset - Prepartum features

Finally, the last experiment evaluates the performance of the classifiers on imbalanced dataset with prepartum features only.

The confusion matrix in Figure 23 shows that all classification algorithms present TP percentage higher than 80% and TN around 80%, except the Random Forest algorithm that presented a lower performance. Regarding the ROC curve and AUC values, Logistic Regression and XGBoost algorithms have better predictive performance considering the AUC value, 0.871 and 0.872, respectively, as depicted in Figure 24).

**Fig 23:**
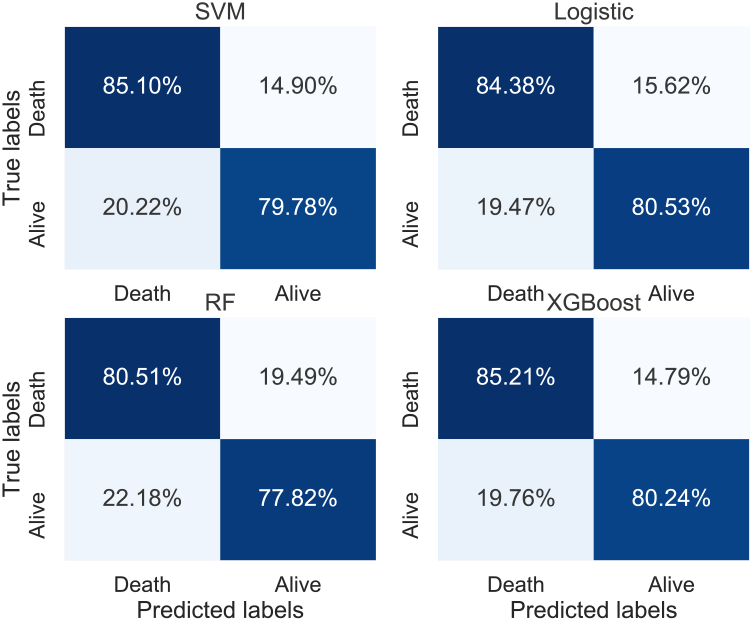
Confusion matrix for balanced dataset with prepartum features.

**Fig 24:**
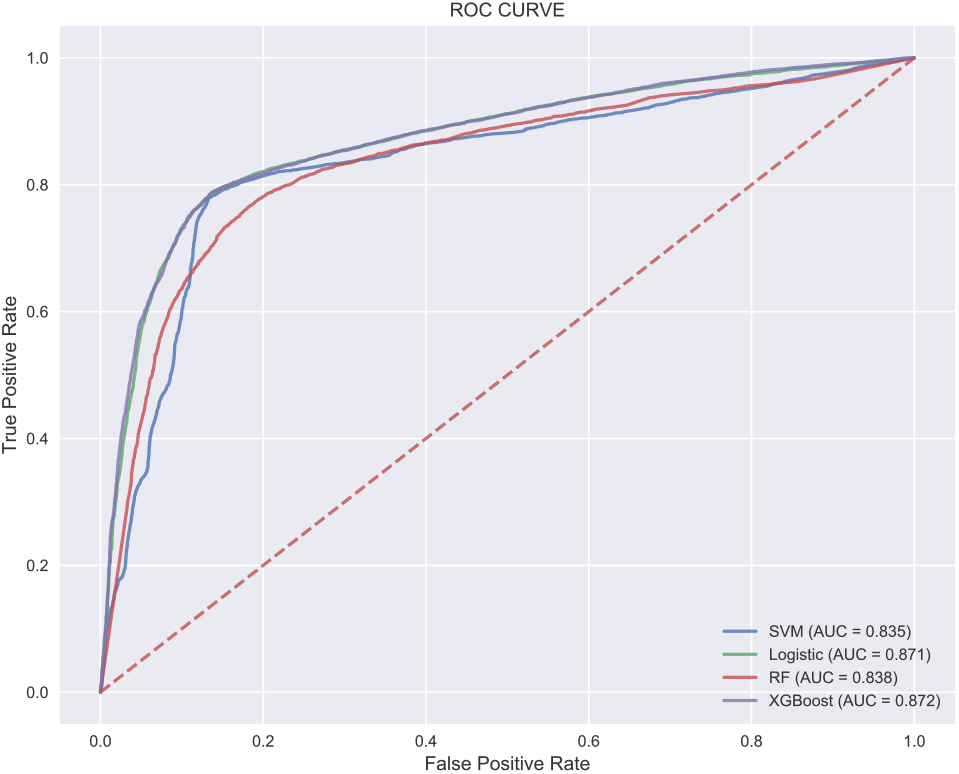
ROC curve for balanced dataset with prepartum features.

Analysing the importance of each prepartum feature on classification models, Figure 25 shows that the number of prenatal appointments and Robson 10-groups predictive features are much more relevant than the others.

**Fig 25:**
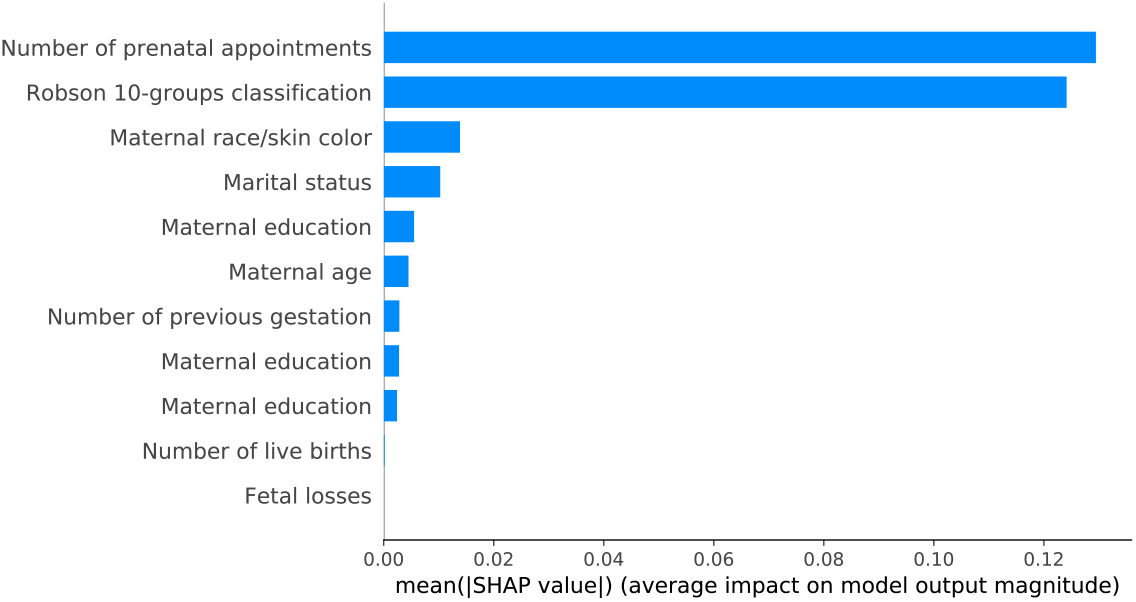
Feature importance for balanced dataset with prepartum features.

## 4 Conclusion and Discussion

This paper presented a methodology to adopt ML algorithms in the task of classifying neonatal mortality using demographic and social-economic features. From public data collected from the Brazilian government, we created a new dataset (SPNeoDeath), comprising more than 1.4*mi* samples for the problem of neonatal mortality.

Approximately one-quarter (28%) of all children worldwide are born with low birth weight [18] and roughly 60 a 80% of all neonatal deaths are associated with this factor [32]. Low birth weight infants are more vulnerable to pulmonary immaturity problems and metabolic disorders, which may cause or aggravate some events that affect them, increasing the risk for mortality. Therefore, low weight and poor ratings in Apgar 1 and 5 are warnings for possible future complications in the child, creating an alert on the risk of this newborn dying in the first days of life. Furthermore, the study of Nascimento *et al*. [32], identified a greater neonatal mortality among premature and low birth weight. Low birth weight is considered a marker of social risk related to precarious socioeconomic conditions and maternal behavior in relation to health care [32].

Features distribution on alive/death samples had been performed, obtaining results which lead us to conclude the high influence on death of the features: number of prenatal appointments, birth weight, apgar indexes and gestational weeks. When applying the feature importance method SHAP, the same features were pointed as being the top 5 influencers of the ML models results. All these results corroborate with Nascimento et. al. [32] and Migoto et. al. [29]. Both low weight and poor ratings in apgar 1 and 5 are warnings for possible future complications in the child, creating an alert on the risk of this newborn dying in the first days of life.

The study of Nascimento et al. [32], identified a greater chance of neonatal mortality among premature, low birth weight and apgar in the 5th minute smaller than 7 (Moderate Score). Low birth weight is considered a marker of social risk related to precarious socioeconomic conditions and maternal behavior in relation to health care [32].

Although the marital status of women is associated with the risk of neonatal mortality, it should be noted that this situation is influenced by the socioeconomic conditions of the region, which impact on the peculiarities of care and morbidity and mortality, to be considered by public health policies [29]. In experiment round #4 this feature figures as the 4th most influencing one in the model.

The descriptive analysis performed in Section 2.1.3 shows that newborns who did not die within the first 28 days of life, 90% were born with weight greater than 2,500 grams. On the other hand, 79% of the neonates who died before the twenty-eighth day of life had insufficient weight - below 2,500 grams showing this association with the target feature as demonstrated in the Figure 10.

Another relevant find is that the feature prenatal consultations is among the top features influencing the models as well as gestational week, as expected. The guarantee of quality and properly conducted prenatal care can detect early maternal and fetal diseases, which may reduce the incidence of premature labor in these cases and the occurrence of low birth weight, a feature highly associated with the outcome of neonatal mortality in the first month of life. As presented in the Figure 9, 78% of the mothers of newborns who do not die within the first 28 days of life, undergo 7 or more prenatal consultations.

There is a vast literature that discusses and relates maternal age to neonatal mortality, such as Nascimento et al [32], França and Lansky [11] and Aquino et al. [2]. Age extremes of the mother (less than 19 years old, especially below 15 years old and age above 35 years old) have traditionally been related to the increased risk of neonatal mortality due to its association with prematurity and low birth weight. This relation between maternal age and neonatal death has been explained through biological, socioeconomic and behavioral factors. In the case of younger women, their biological immaturity would lead to a greater frequency of health problems, premature birth and low weight at birth [2]. In contradiction to this, maternal age did not appeared as one of the most influencing features in model results, being “less” important than maternal color, maternal education and marital status.

Robson 10-groups is a measure of cesarean rate assessment and monitoring. This classification distributes women into 10 groups based on five characteristics: early labor (spontaneous, induced or cesarean), gestational age, fetal presentation, number of fetuses and parity (nulliparous, multiparous with and without previous cesarean section) [27]. The classification is done in the sense of trying to identify and reduce the cesarean rate [27]. It is a characteristic that condenses several measures and models classified as an important feature for the prediction of neonatal mortality.

At rounds #2 and #3 the feature congenital malformations appears as the 2nd most influencing feature in the model results. Newborns who presented some form of malformation, whom presented higher gestational weeks have higher risk of mortality. Congenital anomalies are commonly observed in post term pregnancy, that is, a prolongation of pregnancy beyond term [10]. Higher gestational weeks increase risks of abnormal fetal growth and neonatal death, and this association may be related to the fact that post term fetus may outperform the ability of the placenta to supply nutrients and it’s a risk resulting in malnutrition or asphyxia. Likewise, malformation could be caused by genetic and environmental factors (*e. g*. use of alcohol and tobacco during the pregnancy) [28]. This information is relevant, because some congenital genetic, infectious, or environmental-related anomalies can be prevented through the implementation of public policies and an adequate offer of health services.

Finally, the proposed method is able to deal with missing data, continuous and categorical features, and in the end, classify a new sample according to its chances of dying in the first 28 days of life.

With results exceeding 95% AUC when using XGBoost as a final classifier, the method is able to provide both a death risk response and an interpretation of the result obtained through the use of SHAP values. Between the results across 4 rounds of experiments using 4 different machine learning classifiers with their default parameters, the results point toward expressiveness of features, being feature n ct peso, n st malformacao, n ct apgar5, n ct apgar1, and n tp gestacao respectively the five more expressive as indicated by a specific analysis using SHAP values.

As a decision support tool, this kind of method can be used to help health experts to take decisions if a more intensive care is necessary for newborns. Additionally, from a demographic point of view, studies based on data analysis are valuable to corroborate important statements, once most of the studies are performed in small populations without an expressive statistical sample.

For future research directions, our research group intend to evaluate new methods for dealing with data encoding, such as categorical embeddings, as well as combinations between different classifiers in order to increase positive class (death) accuracy, for the occurrence of false negatives that is a very problematic issue on methods related with health.

Taking advantage of many state-of-the-art algorithms, our method reaches expressive results. Attained results are not only related to accuracy, but with other metrics applied on health problems as ROC curves and AUC, showing effectiveness and efficiency of the proposed method to classify samples according to the risk of death or not. Features used as model input reflect the socioeconomic characteristics of the mother, reproductive history, prenatal care and related characteristics presented by the newborn at birth.

To the best of our knowledge, no other previous works have been proposed with the usage of this dataset along with machine learning algorithms, making it the first of it’s kind in Brazil. Our hypothesis was that neonatal mortality is a complex phenomenon, involving interactions of several characteristics and requiring a large volume of data for its full understanding. In this sense, we believe that traditional regression models may not be sufficient to understand this problem, since the assumptions of parametric modeling are unrealistic for investigations of this nature.

## Data Availability

Dataset will be made public after paper been published in a journal.

## Footnotes

1 In order to allow the reproducibility the of proposed method, along with the comparison with another techniques, source code will be made public on GitHub under paper acceptance.

2 For this paper, a FN answer, when a death sample is classified as alive, it is much more problematic than a FP, when an alive sample is classified as death.

